# Sex-Related Outcomes of Transcatheter Aortic Valve Implantation with Self-Expanding or Balloon-Expandable Valves: Insights from the OPERA-TAVI Registry

**DOI:** 10.1101/2023.09.05.23295098

**Authors:** Marianna Adamo, Luca Branca, Elisa Pezzola, Francesco Saia, Thomas Pilgrim, Mohamed Abdel-Wahab, Philippe Garot, Caterina Gandolfo, Claudia Fiorina, Sofia Sammartino, Azeem Latib, Ignacio Amat Santos, Darren Mylotte, Federico De Marco, Ole De Backer, Luis Nombela Franco, Mariama Akodad, Flavio Luciano Ribichini, Francesco Bedogni, Giulia Laterra, Alessandro Mazzapicchi, Daijiro Tomii, Pietro Laforgia, Stefano Cannata, Andrea Scotti, Simone Fezzi, Enrico Criscione, Enrico Poletti, Mattia Mazzucca, Mattia Lunardi, Andrea Mainardi, Stefano Andreaggi, Angelo Quagliana, Nicholas Montarello, Breda Hennessey, Matias Mon-Noboa, David Meier, Carmelo Sgroi, Claudia Maria Reddavid, Orazio Strazzieri, Silvia Crescenzia Motta, Valentina Frittitta, Elena Dipietro, Alessandro Comis, Chiara Melfa, Mariachiara Calì, Holger Thiele, John G. Webb, Lars Sondergaard, Corrado Tamburino, Marco Metra, Giuliano Costa, Marco Barbanti

## Abstract

**Background:** Evidence regarding sex-related differences in response to transcatheter aortic valve implantation according to the valve type is lacking. This study sought to evaluate the impact of sex on the treatment effect of Evolut-PRO/PRO+ (PRO) or Sapien 3 Ultra (ULTRA) devices on clinical outcomes.

**Methods:** Comparative Analysis of Evolut PRO vs Sapien 3 Ultra Valves for Transfemoral Transcatheter Aortic Valve Implantation (OPERA-TAVI) is a multicenter multinational registry including patients undergoing latest-iteration PRO or ULTRA implantation. Overall, 1174 out of 1897 patients were matched based on valve type and compared according to sex, while 470 males and 630 females were matched and compared according to valve type. Thirty-day and 1-year outcomes were evaluated.

**Results:** In both PRO and ULTRA group, males had a higher comorbidity burden, while females had smaller aortic root. Both 30-day (device success [DS], early safety outcome, permanent pacemaker implantation [PPI], patient-prosthesis mismatch [PPM], paravalvular regurgitation [PVR], bleedings, vascular complications, and all-cause death) and 1-year outcomes (all-cause death, stroke and heart failure hospitalization) did not differ according to sex in both valve groups. However, male sex decreased the likelihood of 30-day DS with ULTRA versus PRO (p for interaction 0.047). A higher risk of 30-day PPI and 1-year stroke, and a lower risk of PPM was observed in PRO versus ULTRA, regardless of sex. No other differences were noted.

**Conclusions:** Sex did not modify the treatment effect of PRO versus ULTRA on clinical outcomes, with the exception of 30-day DS that was decreased in males (versus females) receiving ULTRA (versus PRO).

## INTRODUCTION

Sex-differences in the pathophysiology of aortic stenosis (AS) are known. (1-4) As compared to men, women have smaller aortic root, lower amount of calcium and higher amount of fibrosis on the aortic valve, as well as more concentric left ventricular hypertrophy.(1-3) These anatomic features might impact on the response to transcatheter aortic valve implantation (TAVI) influencing the risk of complications, i.e. patient prosthesis mismatch (PPM), coronary occlusion, paravalvular regurgitation (PVR), permanent pacemaker implantation (PPI), suicide ventricle, vascular complications, bleedings, and stroke, which were found to be differently distributed according to sex (5-9). Possible sex-related differences in 1-year outcomes have been poorly explored. A greater mortality benefit in women as compared to men has been reported in a large patient-level meta-analysis including patients undergoing TAVI, with women also having a lower comorbidity burden. (5)

As well-known, TAVI outcomes can also be influenced by the transcatheter aortic valve (TAV) type, i.e. self-expanding (SE) versus balloon expandable (BE), with SE leading to higher rates of PPI and lower rates of PPM, as compared to BE. (10-12)

However, evidence regarding sex-related differences in the early and mid-term response to TAVI according to the TAV type is lacking. In a female population receiving TAVI, no differences were observed in clinical outcomes according to TAV type (SE versus BE) up to 1-year follow-up, except for PPI rate that was higher with SE device. However, no information regarding the male counterpart is available. (13)

Thus, the aim of this study was to investigate sex-related differences in 30-day and 1-year outcomes in patients undergoing TAVI with new-generation SE (Evolut PRO/PRO+) or BE (Sapien 3 ULTRA) valve.

## METHODS

### Population

OPERA-TAVI (Comparative Analysis of Evolut PRO vs Sapien 3 Ultra Valves for Transfemoral Transcatheter Aortic Valve Implantation) is a registry including data from patients undergoing transfemoral TAVI with Evolut PRO/PRO+ (PRO) or Sapien 3 ULTRA (ULTRA) devices at 14 centres from Europe and North America, between September 2017 and January 2022. Details regarding the registry design have been previously reported. (12) Briefly, exclusion criteria for the analysis were as following: patients who were not eligible for both PRO or ULTRA devices indifferently according to the manufacturers’ instruction for annular dimensions, TAVI in pure aortic valve regurgitation and in degenerated surgical bio-prothesis, patients without pre-procedural CT and 1-year follow up data.

For the purpose of the present analysis the population was stratified according to sex and valve type: males versus females among PRO and ULTRA groups, and PRO versus ULTRA among males and females.

### Outcomes

Both 30-day and 1-year outcomes, defined according to VARC-3 recommendations (14), were assessed. Outcomes of interest at 30-day were: device success, early safety outcome, rate of PPI, PPM, PVR, bleedings, vascular complications, stroke and all-cause death.

Device success was defined as: 1) technical success; 2) 30-day freedom from mortality; 3) 30-day freedom from surgery or intervention related to the device or a major vascular, access-related, or cardiac structural complication; and 4) intended performance of the valve (mean gradient <20 mm Hg, peak velocity <3 m/s, Doppler velocity index ≥ 0.25, and less than moderate aortic regurgitation). The early safety outcome was defined as: 1) freedom from all-cause mortality; 2) freedom from all stroke; 3) freedom from VARC type 2 to 4 bleeding; 4) freedom from major vascular, access-related, or cardiac structural complications; 5) freedom from acute kidney injury stage 3 or 4; 6) freedom from moderate or severe aortic regurgitation; 7) freedom from PPI caused by procedure related conduction abnormalities; and 8) freedom from surgery or intervention related to the device at 30 days.

Outcomes of interest at 1-year were: composite of all-cause death, disabling stroke and heart failure hospitalization (HFH), and all-cause death, disabling stroke and HFH as separate outcomes.

### Statistical analysis

Categoric variables are reported as counts and percentages. Continuous variables are reported as median (IQR). Continuous variables were compared with the Student’s t-test or Mann-Whitney U test for paired samples, and categoric variables were compared with chi-square statistics or using the Fisher exact test for paired samples as appropriate.

To account for the nonrandomized design of our study, adjustment with propensity score matching (PSM) was used. The propensity score was estimated using a logistic regression model according to a non-parsimonious approach. Variables included in the PSM were sex, age, body mass index, diabetes, hypertension, peripheral artery disease, chronic obstructive pulmonary disease, renal failure (defined as estimated glomerular filtration rate <30 mL/min/1.73 m2), prior coronary artery bypass grafting, prior myocardial infarction, prior stroke, prior pacemaker implantation, New York Heart Association functional class (NYHA), coronary artery disease, atrial fibrillation, baseline right bundle branch block, Society of Thoracic Surgeons mortality score, left ventricular ejection fraction, transaortic mean gradient, leaflet and left ventricular outflow tract calcification, bicuspid aortic valve, horizontal aorta, and area/perimeter-derived aortic annulus diameter <23 mm assessed at the preprocedural CT analysis. One-to-one PSM with the nearest neighbour method with a calliper width of 0.1, the SD of propensity score logit, was used in the overall population for the sex-based analysis (males versus females in PRO and ULTRA) and in males and females considered as separate populations to compare PRO and ULTRA matched groups.

The Kaplan-Meier method and the log-rank test were used to evaluate the first occurrence of 1-year clinical outcomes in males versus females in the PRO and ULTRA group and in PRO versus ULTRA in males and females. Logistic regression analysis and Cox proportional hazards regression analysis were performed to assess the risk of 30-day and 1-year outcomes respectively, in PRO versus ULTRA by sex subgroups.

All statistical tests were performed 2-tailed, and a P value <0.05 was considered the threshold for statistical significance (P < 0.10 was the threshold for interaction tests). All statistical analyses were performed with R software version 3.6.3 (R Foundation for Statistical Computing).

## RESULTS

Among the 1897 patients included, 1174 were matched according to valve type and compared, into each group (PRO and ULTRA) according to sex (males versus females), while 470 males and 630 females were matched and compared according to valve type (PRO versus ULTRA) **(Figure 1)**.

**Figure 1.**
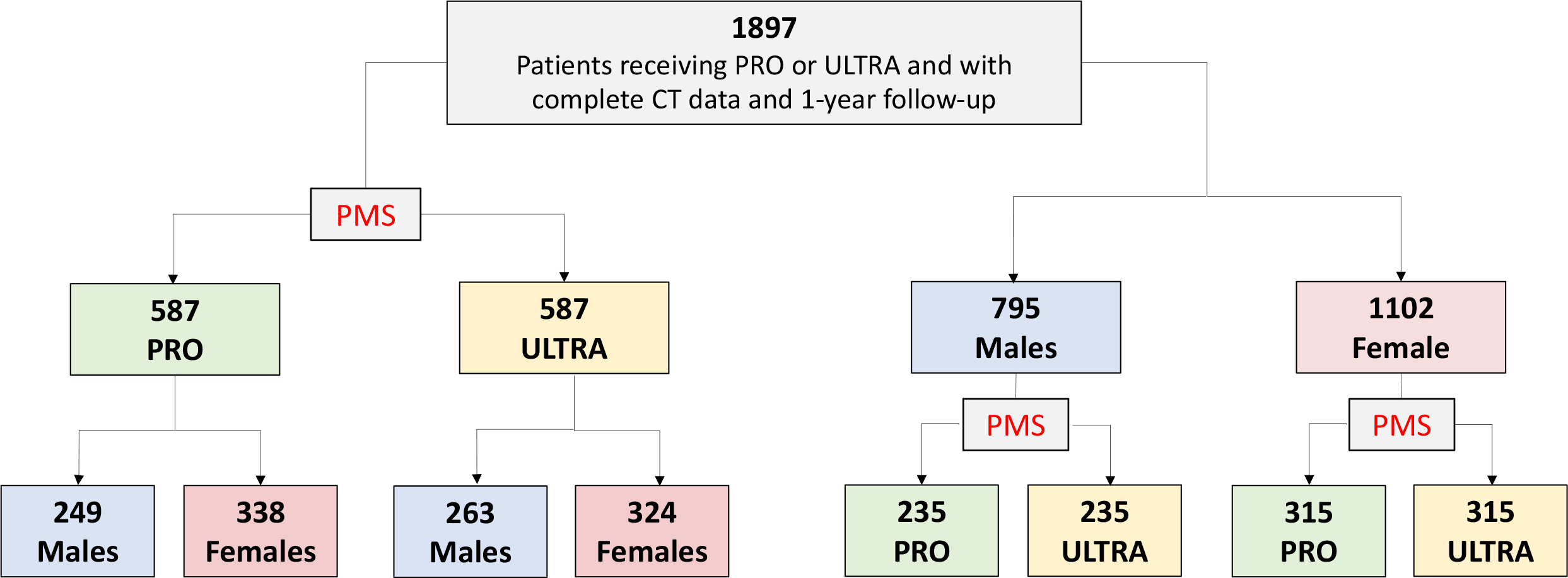
Study Flow Chart. Total number of patients included and number of patients obtained after matching based on valve type in the overall population and in the population stratified by sex

### Baseline characteristics in males versus females by valve type

Among the 587 matched pairs obtained after overall adjustment, 249 (42.4%) males and 338 (57.6%) females were observed in the PRO group, and 263 (44.8%) males and 324 (55.2%) females in the ULTRA group **(Figure 1)**. In both PRO and ULTRA group, males were more likely to have coronary artery disease, higher values of STS score and larger aortic valve area (AVA) as compared to females. Notably, a previous pacemaker implantation was more frequently observed in males versus females only in the PRO group. Aortic root dimensions (annulus, coronary height, sinus of Valsalva, sinus tubular junction) were smaller in females versus males in both PRO and ULTRA group, while valvular calcifications are more frequent in males only in the PRO group **(Table 1)**.

**Table 1.** Baseline clinical, echocardiographic and computed tomography characteristics in males versus females by valve type.

|  |  |  |
| --- | --- | --- |
|  | EVOLUT PRO/PRO+ | SAPIEN ULTRA |

|  | Male<br>(N = 249) | Female<br>(N = 338) | P<br>value | Male<br>(N = 263) | Female<br>(N = 324) | P<br>value |
| --- | --- | --- | --- | --- | --- | --- |
| Age, Y | 82.00<br>(77.00 - 85.22) | 83.00<br>(78.89 - 86.02) | 0.032 | 81.64<br>(76.91 - 86.37) | 82.39<br>(77.86 - 86.30) | 0.505 |
| BMI, Kg/m<sup>2</sup> | 26.22<br>(23.70 - 29.00) | 26.33<br>(23.02 - 31.20) | 0.406 | 26.35<br>(23.88 - 29.39) | 26.45<br>(23.45 - 30.54) | 0.412 |
| Hypertension | 213 (85.5) | 290 (85.8) | 1.000 | 224 (85.2) | 277 (85.5) | 0.907 |
| Diabetes Mellitus | 78 (31.3) | 90 (26.6) | 0.230 | 85 (32.3) | 79 (24.4) | 0.034 |
| CKD | 25 (10.0) | 37 (10.9) | 0.361 | 17 (6.5) | 35 (10.8) | 0.180 |
| CAD | 112 (45.0) | 108 (32.0) | 0.001 | 130 (49.4) | 107 (33.0) | <0.001 |
| Prior MI | 37 (14.9) | 24 (7.1) | 0.004 | 34 (12.9) | 27 (8.3) | 0.089 |
| Prior CABG | 17 (6.8) | 15 (4.4) | 0.269 | 21 (8.0) | 12 (3.7) | 0.030 |
| Prior PM | 28 (11.2) | 19 (5.6) | 0.020 | 23 (8.7) | 22 (6.8) | 0.436 |
| PAD | 36 (14.5) | 39 (11.5) | 0.318 | 36 (13.7) | 41 (12.7) | 0.714 |
| AF | 66 (26.5) | 81 (24.0) | 0.501 | 66 (25.1) | 81 (25.0) | 1.000 |
| Prior Stroke | 29 (11.6) | 28 (8.3) | 0.204 | 40 (15.2) | 24 (7.4) | 0.003 |
| COPD | 33 (13.3) | 34 (10.1) | 0.102 | 29 (11.0) | 38 (11.7) | 0.949 |
| NYHA functional class |  |  |  |  |  |  |
| I | 6 (2.4) | 9 (2.7) | 0.182 | 9 (3.4) | 14 (4.3) | 0.099 |
| II | 89 (35.7) | 113 (33.4) |  | 111 (42.2) | 112 (34.6) |  |
| III | 143 (57.4) | 188 (55.6) |  | 127 (48.3) | 181 (55.9) |  |
| IV | 8 (3.2) | 26 (7.7) |  | 13 (4.9) | 17 (5.2) |  |
| NYHA III-IV | 151 (60.6) | 214 (63.3) | 0.609 | 140 (53.2) | 198 (61.1) | 0.024 |
| Prior RBBB | 20 (8.0) | 20 (5.9) | 0.352 | 25 (9.5) | 23 (7.1) | 0.464 |
| STS mortality score | 2.98<br>(2.04 - 4.29) | 3.41<br>(2.38 - 4.90) | 0.001 | 2.94<br>(1.78 - 4.30) | 3.33<br>(2.14 - 4.89) | 0.005 |
| LVEF | 60.00<br>(55.00 - 64.00) | 60.00<br>(55.00 - 65.00) | 0.281 | 60.00<br>(55.00 - 65.00) | 60.00<br>(53.75 - 65.00) | 0.500 |
| Aortic Peak Gradient, mmHg | 74.00<br>(59.25 - 89.75) | 73.00<br>(59.75 - 87.00) | 0.616 | 70.00<br>(55.25 - 82.00) | 73.00<br>(61.00 - 86.00) | 0.075 |
| Aortic Mean Gradient, mmHg | 44.00<br>(36.00 - 55.00) | 44.00<br>(35.00 - 53.00) | 0.453 | 44.00<br>(35.00 - 51.00) | 44.00<br>(36.00 - 53.00) | 0.221 |
| AVA, cm<sup>2</sup> | 0.72<br>(0.60 - 0.88) | 0.65<br>(0.50 - 0.80) | <0.001 | 0.70<br>(0.60 - 0.85) | 0.70<br>(0.50 - 0.80) | 0.007 |
| Annulus Area CT, mm<sup>2</sup> | 450.00<br>(400.00 - 480.00) | 388.00<br>(351.00 - 438.00) | <0.001 | 439.00<br>(388.00 - 483.00) | 410.90<br>(373.65 - 453.00) | <0.001 |
| Annulus Perimeter CT, mm | 76.25<br>(72.47 - 79.00) | 71.00<br>(67.45 - 75.00) | <0.001 | 76.00<br>(71.30 - 79.15) | 73.00<br>(70.00 - 77.00) | <0.001 |
| LM Height, mm | 14.30<br>(12.75 - 16.85) | 13.00<br>(11.10 - 15.38) | <0.001 | 14.10<br>(12.00 - 16.65) | 13.50<br>(11.90 - 15.20) | 0.023 |
| RCA Height, mm | 16.00<br>(14.00 - 18.70) | 15.30<br>(13.00 - 17.90) | 0.008 | 17.00<br>(14.95 - 19.95) | 15.30<br>(13.00 - 17.50) | <0.001 |
| Moderate to severe leaflet calcification | 178 (72.4) | 208 (62.1) | 0.010 | 171 (66.5) | 211 (65.5) | 0.860 |
| Moderate to severe LVOT calcification | 24 (10.0) | 27 (8.6) | 0.557 | 31 (12.3) | 28 (9.1) | 0.268 |
| STJ mean diameter, mm | 28.40<br>(26.80 - 30.60) | 26.50<br>(24.50 - 29.00) | <0.001 | 29.00<br>(27.30, 30.92) | 27.95<br>(26.00 - 29.70) | <0.001 |
| SoV mean diameter, mm | 32.22<br>(30.08 - 34.63) | 29.27<br>(27.80 - 31.30) | <0.001 | 32.33<br>(30.50, 34.12) | 30.10<br>(28.50 - 32.00) | <0.001 |
| Horizontal aorta | 40 (25.0) | 78 (32.5) | 0.118 | 57 (31.0) | 51 (22.7) | 0.071 |
| Bicuspid aortic valve | 19 (7.7) | 22 (6.5) | 0.625 | 16 (6.1) | 20 (6.2) | 1.000 |

Values are median (IQR) or n (%). BMI, Body Mass Index; CKD, Chronic Kidney Disease; CAD, Coronary Artery Disease; MI, Myocardial Infarction; CABG, Coronary Artery Bypass Graft; PM, Pace-Maker; PAD, Periferic Artery Disease; AF, Atrial Fibrillation; COPD, Chronic Obstructive Pulmonary Disease; NYHA, New York Heart Association; RBBB, Right Bundle Branch Block; STS Score, Society of Thoracic Surgeons Score; LVEF, Left Ventricular Ejection Fraction; AVA, Aortic Valve Area; CT, Computed Tomography; LM, Left Main; RCA, Right Coronary Artery; LVOT, Left Ventricular Outflow Tract; STJ, Sinotubular Junction; SoV, Sinus of Valsalva.

### Baseline characteristics in PRO versus ULTRA by sex

After adjustment in males and females taken separately, 235 and 315 matched pairs treated with PRO or ULTRA devices were obtained in the male and female cohorts, respectively **(Figure 1)**. Baseline demographic and clinical characteristics were well balanced between the two study groups in both cohorts with all standardized mean differences below 10% except for AVA that was smaller in females receiving PRO versus ULTRA device. Regarding preprocedural CT analysis, both males and females treated with PRO versus ULTRA showed smaller sinus tubular junction diameters; females also had smaller annular perimeters **(Table 2)**.

**Table 2.** Baseline clinical, echocardiographic and computed tomography characteristics in Evolut PRO/PRO+ versus Sapien 3 ULTRA by sex.

|  | **PRO**<br>**(N = 235)** | **ULTRA**<br>**(N = 235)** | **P**<br>**value** | **PRO**<br>**(N = 315)** | **ULTRA**<br>**(N = 315)** | **P**<br>**value** |
| --- | --- | --- | --- | --- | --- | --- |
| Age, Y | 82.00<br>(77.00 - 85.00) | 81.77<br>(77.00 - 86.45) | 0.608 | 82.00<br>(78.00 - 85.44) | 82.04<br>(77.93 - 86.32) | 0.630 |
| BMI, Kg/m<sup>2</sup> | 26.37<br>(23.99 - 29.28) | 26.31<br>(23.62 - 29.33) | 0.661 | 26.17<br>(22.90 - 30.95) | 26.50<br>(23.44 - 30.11) | 0.642 |
| Hypertension | 199 (84.7) | 204 (86.8) | 0.598 | 278 (88.3) | 277 (87.9) | 1.000 |
| Diabetes Mellitus | 77 (32.8) | 75 (31.9) | 0.921 | 79 (25.1) | 84 (26.7) | 0.716 |
| CKD | 20 (8.5) | 20 (8.5) | 1.000 | 39 (12.4) | 34 (10.8) | 0.759 |
| CAD | 108 (46.0) | 114 (48.5) | 0.644 | 104 (33.0) | 113 (35.9) | 0.502 |
| Prior CABG | 20 (8.5) | 21 (8.9) | 1.000 | 15 (4.8) | 15 (4.8) | 1.000 |
| Prior PM | 24 (10.2) | 27 (11.5) | 0.767 | 24 (7.6) | 20 (6.3) | 0.640 |
| PAD | 40 (17.0) | 35 (14.9) | 0.615 | 34 (10.8) | 42 (13.3) | 0.392 |
| AF | 63 (26.8) | 65 (27.7) | 0.918 | 78 (24.8) | 77 (24.4) | 1.000 |
| Prior Stroke | 26 (11.1) | 32 (13.6) | 0.483 | 23 (7.3) | 23 (7.3) | 1.000 |
| COPD | 25 (10.6) | 32 (13.6) | 0.326 | 34 (10.8) | 33 (10.5) | 1.000 |
| NYHA functional class |  |  |  |  |  |  |
| I | 9 (3.8) | 8 (3.4) | 0.746 | 10 (3.2) | 12 (3.8) | 0.408 |
| II | 87 (37.0) | 90 (38.3) |  | 102 (32.4) | 109 (34.6) |  |
| III | 128 (54.5) | 120 (51.0) |  | 178 (56.5) | 179 (56.8) |  |
| IV | 9 (3.8) | 15 (6.4) |  | 25 (7.9) | 15 (4.8) |  |
| NYHA III-IV | 137 (58.3) | 135 (57.4) | 0.972 | 203 (64.4) | 194 (61.6) | 0.509 |
| Prior RBBB | 17 (7.2) | 21 (8.9) | 0.741 | 16 (5.1) | 18 (5.7) | 0.918 |
| STS mortality score | 3.13<br>(1.98 - 4.30) | 2.90<br>(1.74 - 4.07) | 0.208 | 3.37<br>(2.38 - 5.00) | 3.36<br>(2.16 - 4.93) | 0.361 |
| LVEF | 60.00<br>(55.00 - 64.00) | 60.00<br>(53.00 - 65.00) | 0.888 | 60.00<br>(55.00 - 65.00) | 60.00<br>(54.00 - 65.00) | 0.728 |
| Aortic Peak Gradient, mmHg | 73.00<br>(59.00 - 88.00) | 70.00<br>(59.75 - 82.00) | 0.263 | 73.00<br>(60.00 - 87.50) | 72.50<br>(60.75 - 86.00) | 0.894 |
| Aortic Mean Gradient, mmHg | 44.00<br>(36.00 - 55.00) | 44.00 (36.00 -<br>52.00) | 0.337 | 45.00<br>(36.00 - 54.00) | 44.00<br>(35.50 - 52.00) | 0.566 |
| AVA, cm<sup>2</sup> | 0.70<br>(0.60 - 0.85) | 0.70<br>(0.60 - 0.83) | 0.204 | 0.60<br>(0.50 - 0.78) | 0.70<br>(0.51 - 0.80) | 0.023 |
| Annulus Area CT, mm<sup>2</sup> | 447.00<br>(400.00 - 480.00) | 444.00<br>(399.50 - 489.25) | 0.525 | 390.00<br>(352.00 - 450.00) | 410.00<br>(373.00 - 451.75) | 0.003 |
| Annulus Perimeter CT, mm | 76.00<br>(72.40 - 79.00) | 76.60<br>(72.62 - 80.00) | 0.159 | 71.00<br>(68.00 - 75.90) | 73.00<br>(70.00 - 76.93) | <0.001 |
| LM Height, mm | 14.30<br>(12.60 - 17.00) | 14.00<br>(12.00 - 16.50) | 0.247 | 13.50<br>(11.60 - 15.40) | 13.50<br>(11.85 - 15.40) | 0.594 |
| RCA Height, mm | 16.20<br>(14.00 - 18.40) | 17.00<br>(14.93 - 19.90) | 0.010 | 15.20<br>(13.00 - 18.00) | 15.30<br>(13.00 - 17.55) | 0.690 |
| Moderate to severe leaflet<br>calcification | 162 (68.9) | 166 (70.6) | 0.842 | 202 (64.1) | 200 (63.5) | 0.975 |
| Moderate to severe LVOT<br>calcification | 26 (11.1) | 28 (11.9) | 0.922 | 29 (9.2) | 28 (8.9) | 1.000 |
| STJ mean diameter, mm | 28.40<br>(26.90 - 30.68) | 29.00<br>(27.70 - 30.95) | 0.043 | 27.00<br>(25.00 - 29.00) | 28.00<br>(26.00 - 29.85) | <0.001 |
| SoV mean diameter, mm | 32.23<br>(30.28 - 34.72) | 32.53<br>(30.58 - 34.97) | 0.346 | 29.93<br>(28.00 - 31.92) | 30.00<br>(28.50 - 32.02) | 0.082 |
| Horizontal aorta | 39 (16.6) | 45 (19.1) | 0.724 | 57 (18.1) | 55 (17.5) | 0.908 |
| Bicuspid aortic valve | 16 (6.8) | 17 (7.2) | 0.859 | 20 (6.3) | 18 (5.7) | 0.867 |

Values are median (IQR) or n (%). BMI, Body Mass Index; CKD, Chronic Kidney Disease; CAD, Coronary Artery Disease; CABG, Coronary Artery Bypass Graft; PM, Pace-Maker; PAD, Periferic Artery Disease; AF, Atrial Fibrillation; COPD, Chronic Obstructive Pulmonary Disease; NYHA, New York Heart Association; NA, Not Assessed; RBBB, Right Bundle Branch Block; STS Score, Society of Thoracic Surgeons Score; LVEF, Left Ventricular Ejection Fraction; AVA, Aortic Valve Area; CT, Computed Tomography; LM, Left Main; RCA, Right Coronary Artery; LVOT, Left Ventricular Outflow Tract; STJ, Sinotubular Junction; SoV, Sinus of Valsalva.

### Procedural details

Procedural data are reported in **Supplementary Tables 1 and 2**.

As expected, females received smaller valve size compared with males, in both PRO and ULTRA group. Only in the PRO group, males as compared to females, more frequently received predilatation (49% vs. 38%; p=0.015), concomitant percutaneous coronary interventions (6.8%% vs. 2.7%; p=0.024) and higher contrast medium amount (130 [85 - 172] ml vs. 110 [84 - 151] ml; p=0.002)

Rate of pre and postdilatation as well as contrast medium amount were higher in PRO versus ULTRA in both sexes.

### Early outcomes

No differences in the rate of in-hospital complications and 30-day clinical outcomes and echocardiographic data were observed in males versus females in both PRO and ULTRA groups **(Supplementary Table 3 and 4)**.

Regression analysis showed a higher risk of 30-day PPI and a lower risk of any 30-day PPM in PRO versus ULTRA in both males and females. Moreover, sex had a significant impact on the treatment effect of PRO versus ULTRA on 30-day device success, with ULTRA being associated with a lower performance as compared to PRO only in males **(Figure 2)**. No differences in the risk of 30-day all-cause death, any bleedings, vascular complications and moderate/severe PVR were observed in PRO versus ULTRA group, also after sex-matched analysis **(Figure 2)**. The rate of stroke and HF hospitalizations was very low with no significant differences between groups **(Supplementary Table 5)**. Notably, in both females and males, the rate of any PVR was higher in PRO versus ULTRA, while the rate of severe PPM was similar in PRO and ULTRA **(Supplementary Table 6)**. Also, the rate of major bleedings and new onset atrial fibrillation was higher only in females receiving PRO versus ULTRA **(Supplementary Table 5)**, but sex did not modify the impact of valve type on this outcome (p for interaction 0.453).

**Figure 2.**
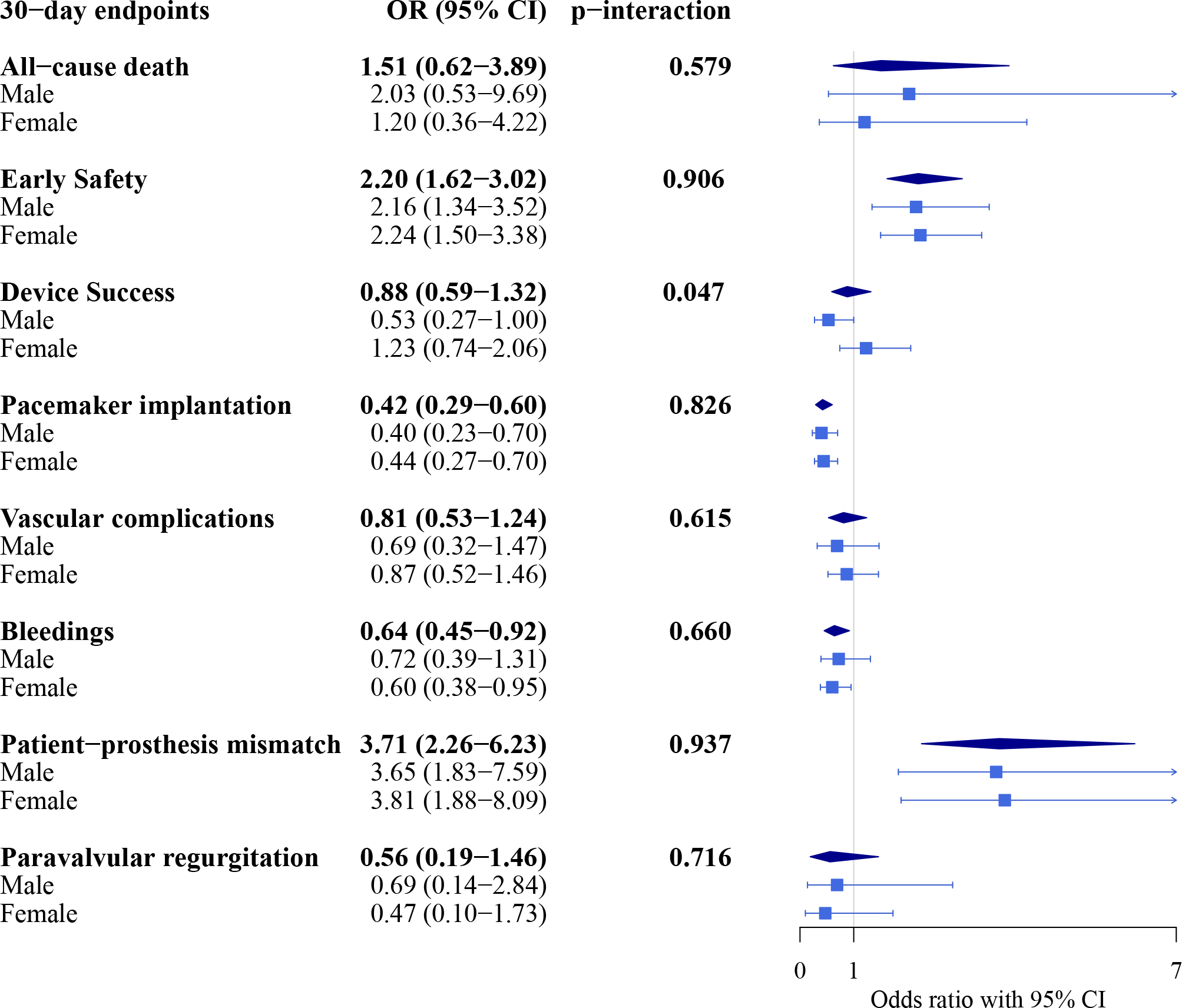
Thirty-day clinical outcomes. Odds ratio (OR) and 95% confidence interval (CI) for all the 30-day outcomes in PRO versus ULTRA in the overall population and in the two subgroups based on sex. P for interaction is reported for all 30-day outcomes. OR>1 indicates that PRO is better and OR<1 indicates that ULTRA is better, except for early safety and device success where OR>1 indicates that ULTRA is better and OR<1 indicates that PRO is better.

### One-year outcomes

Cumulative incidence of all-cause death, HFH and composite endpoint, including all-cause death, HFH and disabling stroke, did not differ significantly either in males versus females in both PRO and ULTRA groups (all-cause death: males 9.2% vs. females 10.1% in PRO and males 11.4% vs. females 9.9% in ULTRA; HFH: males 3.4% vs. females 2.8% in PRO and males 1.6% vs. females 2.9% in ULTRA; composite endpoint: males 9.6% vs. females 10.7% in PRO and males 11.4% vs. females 9.9% in ULTRA) **(Supplementary Figure 1-4)** or in PRO versus ULTRA in both sexes (all-cause death: PRO 9.2% vs. ULTRA 9.8% in females and PRO 9.8% vs. ULTRA 12.8% in males; HFH: PRO 2.7% vs. ULTRA 3.7% in females and PRO 3.1% vs. ULTRA 1.8% in males; composite endpoint: PRO 9.8% vs. ULTRA 9.8% in females and PRO 10.2% vs. ULTRA 9.8% in males) **(Supplementary Figure 5-7)**. On the other hand, the cumulative incidence of disabling stroke was similar in males and females in both PRO and ULTRA groups (males 2.9% vs. females 2.4% in PRO and males 0.4% vs. females 0.3% in ULTRA), but higher in PRO versus ULTRA in both sexes (PRO 2.6% vs. ULTRA 0% in females and PRO 4.4% vs. ULTRA 0.9% in males) **(Supplementary Figure 8)**. Notably, sex did not impact the treatment effect of PRO versus ULTRA on 1-year outcomes **(Figure 3)**.

**Figure 3.**
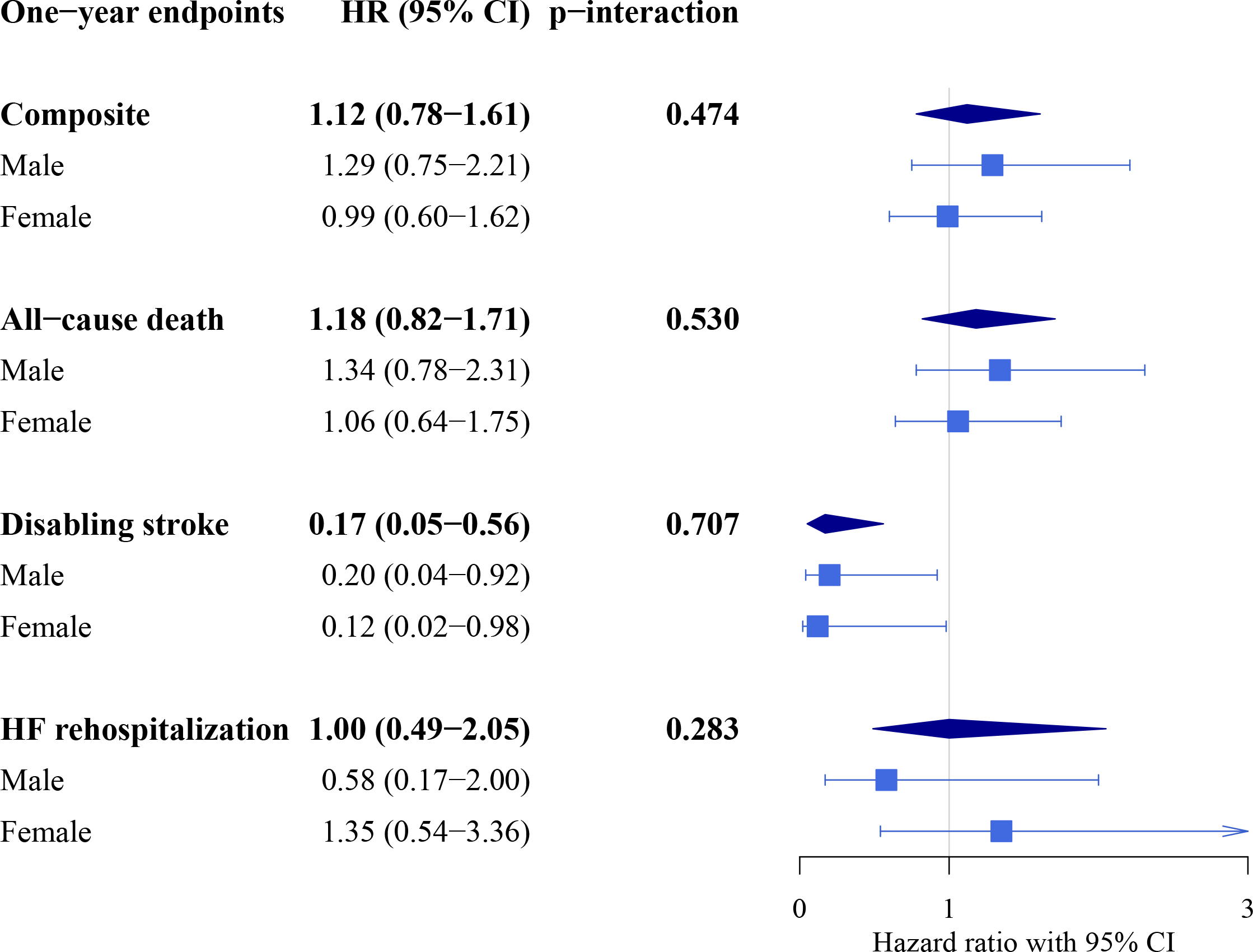
One-year clinical outcomes. Hazards ratio (HR) and 95% confidence interval (CI) for all the 1-year outcomes in PRO versus ULTRA in the overall population and in the two subgroups based on sex. P for interaction is reported for all 1-year outcomes. HR>1 indicates that PRO is better and HR<1 indicates that ULTRA is better.

At 1-year echocardiographic evaluation no differences were noted between females and males in both PRO and ULTRA group **(Supplementary Table 7)**. On the other hand, mean gradients were lower in PRO versus ULTRA in both males and females (PRO 8 (6-12) mmHg vs. ULTRA 11 (8-14) mmHg (p=0.001) in males and PRO 8 (6-12) mmHg vs. ULTRA 12 (8-15) mmHg (p<0.001) in females), while no differences were observed in terms of any and moderate/severe PVR **(Supplementary Table 8)**.

## DISCUSSION

The main finding of the present analysis is that sex does not affect the impact of valve type on 30-day and 1-year outcomes after TAVI despite several sex-differences in baseline presentation and procedural features, with the exception of 30-day device success that was lower in males receiving Sapien 3 Ultra versus Evolut PRO.

Sex-differences in pathophysiology of AS have been previously reported. (1-3) Women have a lower expression of specific MMPs and types of collagens, resulting in a more prevalent valvular fibrosis. On the other hand, the amount of valvular calcification is higher in men. Moreover, women with AS present more concentric left ventricular hypertrophy and smaller cavities than men, smaller aortic annuli and smaller peripheral vessels. (1-3) Sex-differences in the response to AS treatment have also been described. Women seems to benefit more from TAVI than surgery as compared to man. (15) Also, in TAVI setting, women seem to have different outcomes than men. (5-9)

A higher rate of vascular complications and bleedings after TAVI has been reported in females versus males ascribed to smaller and fragile peripheral vessels. (5-9,16,17) In the OPERA-TAVI registry no differences in vascular complications and any bleedings were noted in females versus males in both TAV groups as well as in PRO versus ULTRA in both sexes. Of note, both absolute and relative rates of vascular complications are lower in OPERA than in previous studies. (5,6) The former due to the smaller population included, the latter probably due to the new-generation devices use.

Previous data about sex-differences in the rate of stroke after TAVI are contrasting. Some studies reported higher rates of stroke in females versus males (5) potentially due to smaller aortic valve area, higher baseline gradients and consequent higher need for predilatation. Importantly, the rate of SE was higher in females versus males. Other studies have showed similar (6) or lower (17) rates of 30-day stroke in women versus men. In OPERA-TAVI the rate of stroke was too low at 30-day to draw conclusions. However, at 1-year follow-up, patients receiving PRO had a higher rate of stroke as compared to those treated with ULTRA, independently from sex, with similar event rates in males and females.

Post-TAVI permanent pacemaker implantation (PPI) rate was reported to be lower in women than men with lower calcium amount as the main possible explanation to this finding (5,6). Moreover, in a large meta-analysis, the risk of PPI after TAVI was decreased in females despite a more frequent use of SE valves. However, this association was attenuated in females receiving BE valves. (18) In OPERA-TAVI, after matching for TAV type, there were no differences in PPI rate according to sex. On the other hand, as already known, the PRO valve increased the risk of 30-day PPI as compared to the ULTRA device. Importantly the impact of the valve type on PPI risk was independent from sex.

Increased rates of PVR have been reported in males versus females in previous studies ascribed to a large calcium amount. (1,5) In OPERA-TAVI, despite a higher rate of moderate/severe calcification in men versus women only in the PRO group, the rate of moderate/severe PVR did not differ between sexes either in PRO or ULTRA cohort. Of note, males in the PRO group more frequently received predilatation. The risk of moderate/severe PVR was not increased in PRO versus ULTRA in both sexes, while the rate of mild PVR was lower in ULTRA versus PRO regardless of sex.

Data on PPM after TAVI according to sex are limited. In the WIN-TAVI registry, valve sizing and body mass index are two independent predictors of PPM in women. However, PPM seemed not to have an impact on 1-year clinical outcomes. (19) In the TAVI-SMALL registry, where the majority of the patients were females, severe, but not a moderate PPM was associated with poor outcome, and BE device was found to be the strongest predictor of PPM. (20) In the OPERA-TAVI registry, the rate of PPM did not differ significantly in females versus males in both ULTRA and PRO groups, although, in line with available literature, smaller annuli and smaller valve sizing were observed in females. Moreover, the rate of any PPM, but not severe PPM, was higher in patients receiving ULTRA than PRO regardless of sex.

As also observed in previous studies, men undergoing TAVI in OPERA-TAVI had a high comorbidity burden. (5,6) Specifically, they had higher STS score and were more likely to have coronary artery disease, compare with women. Consequently, the rate of concomitant percutaneous coronary intervention and amount of contrast medium were higher in men than women. However, the latter finding was observed only in the PRO group since a concomitant PCI, immediately before TAVI, may be easier than a staged coronary re-access when dealing with a tall-frame supra-annular TAV. (21,22) Importantly, these differences in the procedural approach did not impact on clinical outcomes since both bleedings, acute kidney injury and mortality did not differ according to sex and/or valve type.

Interestingly, 30-day device success was lower with ULTRA versus PRO only in males (p for interaction 0.047). This novel finding can be explained by the numerically lower rate of technical success (i.e. higher rate of annular rupture) observed in males versus females receiving ULTRA, probably due to unfavorable quality and/or distribution of calcium on the aortic root. An additional explanation might be the numerically higher rate of post-procedural aortic gradient ≥20 mmHg, in males versus females receiving ULTRA, with males more frequently receiving 20 mm sizing as compared to females. Notably, despite a slight divergence of the survival curves in favor of females versus males in the ULTRA group and in favor of PRO versus ULTRA in males, no significant differences in 1-year outcomes were observed between groups. Also, the high comorbidity burden observed in males seemed not to impact on 1-year events. Specifically, in contrast with previous findings, (5,23) mortality rate was similar in males and females up to 1-year after TAVI, as well as in PRO and ULTRA regardless of sex.

## LIMITATIONS

Several limitations of this study must be acknowledged. First, this is an observational registry with site-reported data, including outcomes. Indeed, clinical events were not adjudicated by a Central Committee, and echocardiographic data were not evaluated by an independent Core Laboratory.

Second, although PSM adjustments, unmeasured confounders might remain and have potentially affected the results due to the non-randomized nature of the study.

## CONCLUSIONS

In the OPERA-TAVI registry, although several sex-differences in baseline and procedural characteristics, sex had no impact on the treatment effect of Evolut PRO or PRO+ versus Sapien 3 ULTRA on 30-day and 1-year clinical outcomes, except for 30-day device success that was lower with ULTRA versus PRO only in males.

## Data Availability

Data analyzed in this study will be available if needed subject to Dr Barbanti's authorization

